# The impact of COVID-19 and socioeconomic status on psychological distress in cancer patients

**DOI:** 10.1101/2022.11.21.22282580

**Authors:** Elisabeth L. Zeilinger, Matthias Knefel, Carmen Schneckenreiter, Jakob Pietschnig, Simone Lubowitzki, Matthias Unseld, Thorsten Füreder, Rupert Bartsch, Eva K. Masel, Feroniki Adamidis, Lea Kum, Barbara Kiesewetter, Sabine Zöchbauer-Müller, Markus Raderer, Maria T. Krauth, Philipp B Staber, Peter Valent, Alexander Gaiger

## Abstract

**Background:** The COVID-19 pandemic has affected psychological wellbeing in many aspects, but its influence on cancer patients it not yet clear, and studies show mixed results.

**Aims:** We aimed to investigate the impact of the pandemic on psychological symptom burden against the socio-economic background of cancer patients using data from routine assessments before and during the pandemic.

**Methods:** Standardised assessment instruments were applied in *N* = 1,329 patients to screen for symptoms of anxiety, depression, post-traumatic stress, and fatigue from 2018 to 2022. Two MANOVAs with separate ANOVAs and Bonferroni pairwise comparisons as post-hoc tests were computed. First, only time was included as predictor to examine the isolated impact of the pandemic. Second, income level and education level were included as further predictors to additionally test the predictive power of socioeconomic risk factors. All tests were two-sided.

**Results:** Once indicators of socioeconomic status were included in the analysis, the seeming influence of the pandemic became negligible. Only income had a significant impact on all aspects of psychological symptom burden, with patients with low income being highly burdened (partial η^2^ = .01, *p* = .023). The highest mean difference was found for depressive symptoms (*MD* = 0.13, *CI* = [0.07; 0.19], *p* < .001). The pandemic had no further influence on psychological distress.

**Conclusions:** Although the pandemic is a major stressor in many respects, poverty is by far the most important risk factor for psychological symptom burden in cancer outpatients and outweighs the impact of the pandemic.

## 1. Introduction

The COVID-19 pandemic has a significant impact on mental health in various groups of people all over the world (1,2). People with a cancer diagnosis are at particular risk of developing comorbid mental disorders (3), that not only affect quality of life, but also have a significant impact on physical outcomes and survival (4,5).

Psychological distress in cancer patients might have further increased during the pandemic (6). The need for mental health care and counselling was reported to be “skyrocketing” (7). However, studies on mental health problems in cancer patients during the pandemic report mixed results (8). While some studies found elevated mental health problems, including symptoms of depression and anxiety (6,9), others found no increase in psychological symptom burden (10,11). One shortcoming of previous research is the exclusive use of data collected during the pandemic, which cannot be reliably compared with the pre-pandemic period. Furthermore, sociodemographic factors may mediate the mental health response of cancer patients during the COVID-19 pandemic. Recent research found that socioeconomic status (SES) interacts with individual response to both containment measures’ extension and ending (12), and to healthcare seeking behavior (13).

Our research objective was to investigate the impact of the COVID-19 pandemic and of socioeconomic status on psychological symptom burden of cancer outpatients by comparing data routinely collected before and during the pandemic.

## 2. Materials and method

We have followed the STrengthening the Reporting of OBservational studies in Epidemiology (STROBE; 14) guidelines in our reporting.

### 2.1. Materials

Data was collected by means of a questionnaire that is part of the routine assessment in our outpatient clinic. This questionnaire consisted of a sociodemographic profile, including the SES indicators monthly net household income and educational level, and standardized assessment instruments, namely the Hospital Anxiety and Depression Scale (HADS; 15), the Post-Traumatic Symptom Scale (PTSS-10; 16), and a visual analogue scale (VAS) to assess fatigue.

#### 2.1.1. Hospital Anxiety and Depression Scale

The HADS is a 14 item self-report screening tool with seven items each relating to anxiety and depression (15). Psychometric evaluations indicated good results in cancer patients (17). All items are rated on a 4-point Likert scale ranging from zero to three. Two scores can be calculated: the depressive symptoms score (HADS-D) and the anxiety symptoms score (HADS-A). Higher scores represent higher symptom burden. Scores up to seven indicate no depression/anxiety, scores between eight and ten imply a possible anxiety/depressive disorder, and scores higher than ten indicate significant depressive/anxiety symptoms. Internal consistencies (Cronbach α) in the present study sample were high, with α = .86 for the HADS-D, α = .84 for the HADS-A.

#### 2.1.2. Post-Traumatic Symptom Scale

The PTSS-10 is a ten item self-report instrument to assess post-traumatic stress symptoms (PTSS). Each item is rated from zero to three, with higher scores indicating higher symptom burden. A total score higher than 12 implies significant PTSS. Psychometric evaluations indicate that the PTSS-10 is a responsive, valid and reliable screening tool for PTSS (16). Internal consistency in the present study sample was high, with Cronbach α = .86.

#### 2.1.3. Visual Analogue Scale

Fatigue was measured on a one-item VAS ranging from zero to ten. This assessment method was shown to be feasible and valid in cancer patients (18), with even higher sensitivity and reproducibility than a Likert scale (19).

### 2.2. Procedure

Data for this study were assessed at the hematological and oncological outpatient clinic of the Vienna General Hospital. The following inclusion criteria were applied: (1) confirmed diagnosis of cancer or other hematologic neoplasms, (2) age ≥ 18, (3) capacity to consent, (4) sufficient German-language skills. After explanation of the study and written informed consent, patients were handed out questionnaires. The response rate was 78 %. The study was approved by the institutional ethics committee of the Medical University of Vienna (EC Nr: 2255/2016; 1241/2021).

### 2.3. Statistical methods

Two multivariate analyses of covariance (MANOVAs) were computed. For both, dependent variables were the scores of depression, anxiety, post-traumatic symptoms (PTSS), and fatigue. In the first analysis, we investigate solely the impact of the COVID-19 pandemic on psychological burden of cancer patients, with time of assessment included as predictor. We used a reference period of two years prior to the pandemic (Mar 18 to Feb 20). The subsequent time during the pandemic (Mar 20 to Jun 22) was split into seven distinct time periods of four months each. This resulted in eight distinct samples from eight time spans for analysis (see Table 1). In the second analysis, we added two SES indicators as further predictors: highest educational level (primary education / secondary education / post-secondary or tertiary education) and monthly net household income (< 1,300 EUR / 1,300 – 2,200 EUR / > 2,200 EUR). Income levels were chosen based on poverty thresholds in Austria (20).

**Table 1.**
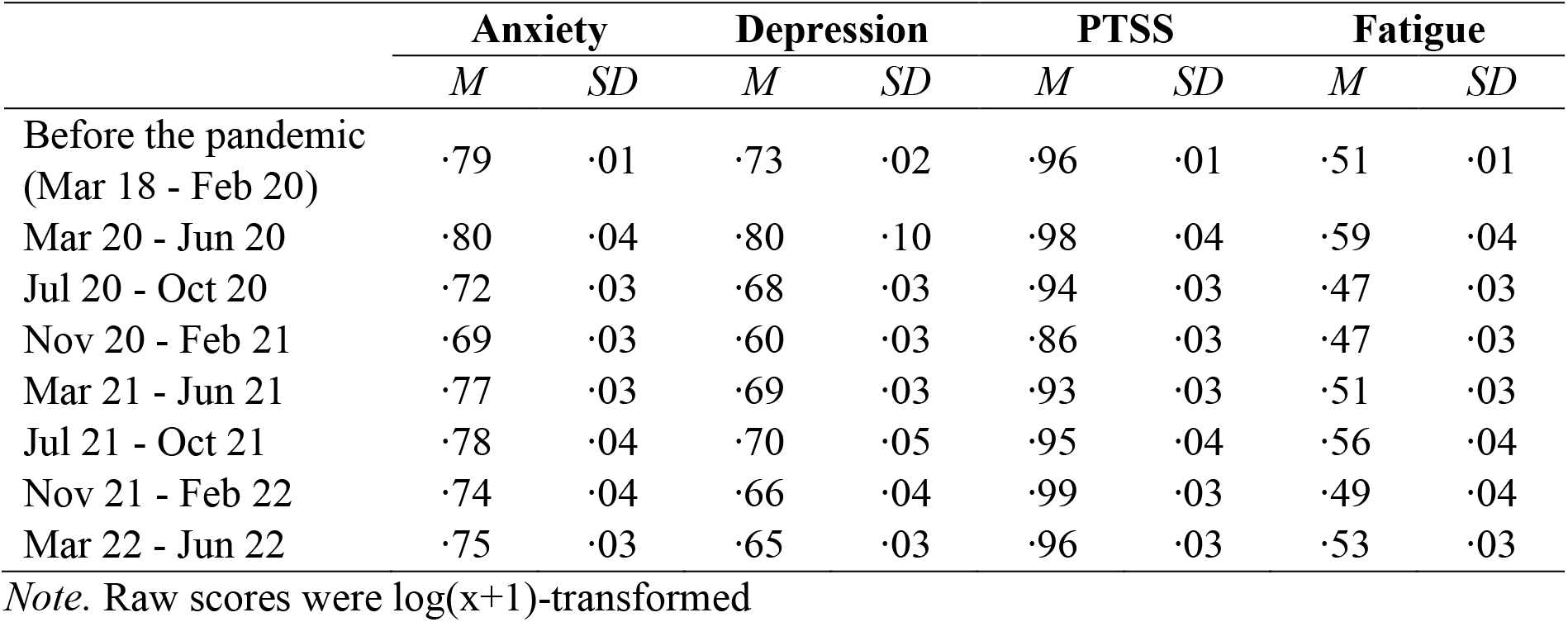
Mean values and standard deviation of the dependent variables upon all time periods

All four dependent variables were log(x+1)-transformed due to high skewness. Transformation was shown to be a robust procedure for right-skewed data in simulation studies (21). Separate ANOVAs and Bonferroni pairwise comparisons were applied as post-hoc tests. Tests were two-sided and Alpha level was set to *p* < .05. No adjustments for multiple testing were made due to an individual interest in each dependent variable and therefore an individual testing approach (22). For ease of graphical interpretation, plotted data in the Figure was z-transformed. This transformation standardizes each scale to a mean of zero and a standard deviation of one, ensuring direct comparability between scales. Analyses were performed using SPSS 28. Due to the nature of this study, randomization or blinding was not applicable. A power analysis was not feasible because a natural sample within the COVID-19 pandemic was analyzed, without the possibility of pre-determining the sample size. The data underlying this article are available in the Open Science Framework (OSF), at https://doi.org/10.17605/OSF.IO/7THFY (23).

**Figure.**
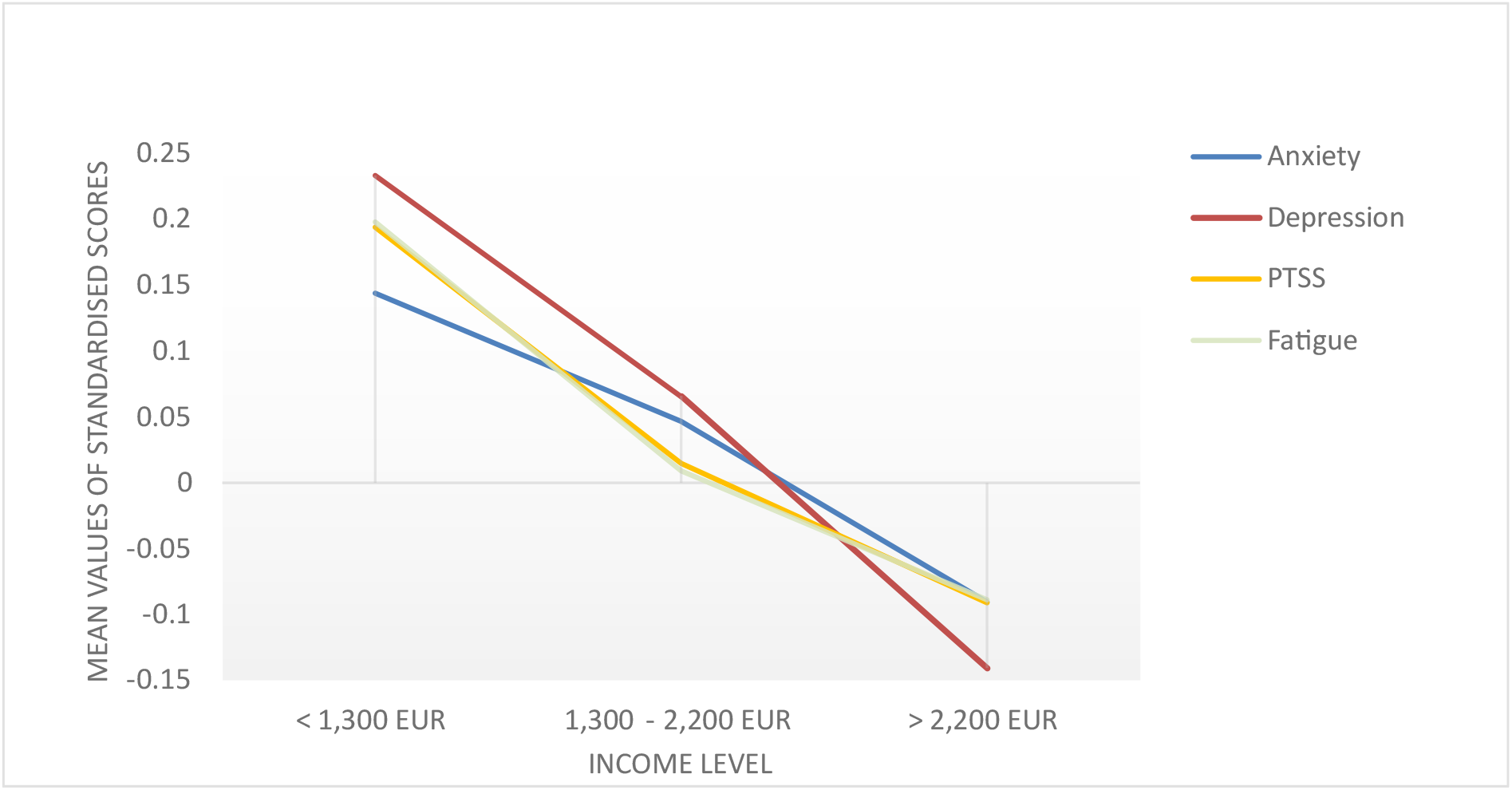
Symptoms of anxiety, depression, PTSS, and fatigue across all income levels. The Figure shows the psychological symptom burden across income levels. Scores have been z-transformed to aid visual interpretation. This transformation standardizes each scale to a mean of zero and a standard deviation of one, ensuring direct comparability between scales.

## 3. Results

The total sample comprised 1,329 outpatients with cancer or other hematologic neoplasms (49.6 % female). Age ranged from 18 to 92 years (*M* = 59, *SD* = 14.14). The sample included 636 patients in the reference timeframe two years prior to the COVID-19 pandemic (from Mar 2018 to Feb 2020), and 693 within the first pandemic years (from Mar 2020 to Jun 2022). Socio-demographic and clinical characteristics of the sample are depicted in Table 2. Scores of anxiety, depression, PTSS, and fatigue across all time periods are shown in Table 1.

**Table 2.** Socio-demographic and clinical characteristics of the sample

| Characteristic | *n* | % |
| --- | --- | --- |
| Gender |  |  |
| Female | 659 | 49.6 |
| Male | 670 | 50.4 |
| Marital status |  |  |
| Single/widowed/divorced | 484 | 36.4 |
| Married/partnered | 845 | 63.6 |
| Educational level |  |  |
| Primary education/apprenticeship | 489 | 36.8 |
| Secondary education | 448 | 33.7 |
| Postsecondary/tertiary education | 392 | 29.5 |
| Monthly net household income |  |  |
| <1,300 Euro | 270 | 20.3 |
| 1,300 – 2,200 Euro | 418 | 31.5 |
| >2,200 Euro | 641 | 48.2 |
| Employment |  |  |
| Employed | 616 | 46.3 |
| Unemployed | 94 | 7.1 |
| Retired | 619 | 46.6 |
| Cancer type |  |  |
| Haematologic cancer/neoplasms | 324 | 24.4 |
| Lung | 167 | 12.6 |
| Breast | 132 | 9.9 |
| Soft tissue | 92 | 6.9 |
| Head and neck | 73 | 5.5 |
| Colon/Rectum | 73 | 5.5 |
| Pancreas | 68 | 5.1 |
| Brain | 64 | 4.8 |
| Stomach/Oesophagus | 51 | 3.8 |
| Kidney/Urinary tract/bladder | 38 | 2.9 |
| Melanoma | 29 | 2.2 |
| Female genital organs | 21 | 1.6 |
| Prostate | 21 | 1.6 |
| Hepatobiliary | 19 | 1.4 |
| Thyroid | 12 | 0.9 |
| Other | 145 | 10.9 |

Note. *N* = 1,329

The first MANOVA included only timespan as predictor and showed a statistically significant difference between the respective time periods on the combined dependent variables (*F*(28, 4753.54) = 1.94, *P* = .002, partial η^2^ = .01, Wilk’s Λ = .96). Post-hoc tests showed a statistically significant difference for anxiety (*p* = .018) and depression (*p* < .001), but not for PTSS (*p* = .106) and fatigue (*p* = .197).

In the second MANOVA we included two SES indicators (income and education level) together with timespan as predictors in the analysis. Statistically significant differences were only found between income levels on the combined dependent variables (*F*(8, 2572) = 0.7, *P* = .023, partial η^2^ = .01, Wilk’s Λ = .99). Timespan was not a significant predictor. Post-hoc ANOVAs showed a statistically significant difference for all four dependent variables related to income. Patients with the lowest income level showed significantly higher symptom burden than patients with the highest income level in all dependent variables (*p*_*s*_ ranging from < .001 to .004). The highest mean difference was found for depressive symptoms (*MD* = 0.13, *CI* = [0.07; 0.19], *p* < .001). Table 3 shows all results. In the Figure, mean symptom burden is plotted across income levels and illustrates the high symptom burden in people with low income.

**Table 3.** Pairwise comparisons of income levels on psychological symptom burden.

|  |  | 1,300 EUR – 2,200 EUR |  |  | > 2,200 EUR |  |  |
| --- | --- | --- | --- | --- | --- | --- | --- |
|  |  | Mean difference | 95 % CI | *P* | Mean difference | 95 % CI | *P* |
| Anxiety | < 1,300 EUR | 0.03 | [-0.03; 0.09] | 0.634 | **0.07** | **[0.02; 0.13]** | **0.004** |
|  | 1,300 EUR – 2,200 EUR | - | - | - | 0.43 | [-0.00; 0.08] | 0.087 |
| Depression | < 1,300 EUR | 0.06 | [-0.01; 0.12] | 0.09 | **0.13** | **[0.07; 0.19]** | **< .001** |
|  | 1,300 EUR – 2,200 EUR | - | - | - | **0.07** | **[0.02; 0.12]** | **0.003** |
| PTSS | < 1,300 EUR | 0.06 | [-0.00; 0.12] | 0.063 | **0.09** | **[0.04; 0.15]** | **< .001** |
|  | 1,300 EUR – 2,200 EUR | - | - | - | 0.03 | [-0.01; 0.08] | 0.275 |
| Fatigue | < 1,300 EUR | **0.06** | **[0.01; 0.12]** | **0.045** | **0.09** | **[0.04; 0.12]** | **< .001** |
|  | 1,300 EUR – 2,200 EUR | - | - | - | 0.03 | [-0.02; 0.08] | 0.357 |

*Note.* For each of the four dependent variables, all three income levels are compared with each other. Significant results are marked in bold. Positive mean differences indicate higher symptom burden in patients with lower income.

## 4. Discussion

The present study investigated the impact of the pandemic and SES indicators on psychological burden of cancer outpatients. We found that the pandemic had no impact once SES indicators were included in the analysis. Lower income was the most significant predictor of psychological distress and this effect was independent of the COVID-19 pandemic. Examining only the influence of time, we could have concluded that the pandemic had a direct influence on psychological outcomes. However, when income and education level were included as predictors in addition to timespan, we found that only income was a significant predictor of higher psychological symptom burden across all dependent variables, i.e. anxiety, depression, PTSS, and fatigue.

The pandemic had a significant impact on health care and cancer care in already undeserved groups, including those with low SES (13,24). However, our data indicate, that it does not have a direct impact on psychological distress. It may well be that the generally heavy psychological symptom burden in cancer patients cannot be further elevated by the pandemic, or that the cancer diagnosis as such is the more significant stressor. Among cancer patients, however, there are and always have been patients who are particularly vulnerable, including people of low SES. We show that poverty is a major cause of higher psychological symptom burden, even during the pandemic.

Our data was collected at the Vienna General Hospital, the largest public oncology care center in Austria. It should be noted that the health care system in Austria provides health insurance to every person, regardless of employment status. Therefore, anyone can access oncology care at our center and our sample is not biased by insurance status.

Given the strong impact of psychiatric comorbidities on overall survival in cancer patients (4), there is a need to provide low-threshold, affordable psychosocial care for patients with low income. We also point out, that research efforts examining the impact of the pandemic would be erroneous if other potential influencing factors were not considered. An exclusive focus on the pandemic could be one aspect contributing to the diverging results of studies on this topic.

### 4.1. Strengths and limitations

One strength of this research lies in the standardized routine assessment of psychological distress before and during the COVID-19 pandemic, which provides the opportunity to reliably compare psychological distress across these time periods. However, participation in this study was voluntary. Therefore, we have no influence on sample characteristics, which can bias our results. This also includes the impact of cancer type. Certain cancer types are associated with low SES and poor health behaviors, such as aggressive ENT tumors, which are associated with the risk factors of alcoholism and smoking. We did not control for cancer type and therefore cannot make assumptions about the impact of the pandemic on patients with specific cancers. However, our sample is an unselected mixed cancer sample found at the largest public outpatient clinic in Austria and therefore has high external validity. Another limitation of this study is the use of screening instruments. Although we cannot make statements on psychiatric diagnoses by means of these instruments, they are well-suited to assess psychological symptom burden.

We are aware, that there are multiple factors influencing psychological well-being, and that we only included some of the most prominent aspects, SES indicators, in the analysis. Further aspects, including physical symptom burden, cancer stage, or treatment phase were not included. Yet, considering sample size and statistical power, it would not have been advisable to include more predictors in the present analysis. As a single-center study, the present work is exploratory in nature and needs to be confirmed in other settings or centers.

### 4.2. Conclusion

Since the pandemic has taken hold of the world, it is omnipresent in every part of life, including health care and research. In many respects this is justified, as this novel situation poses innumerable challenges and obstacles that need to be addressed. However, the pandemic is not the only threat that impacts health, health care, psychological wellbeing, and survival. While we are trying to find ways to support patients during the pandemic, which is undoubtedly an essential task, it may also be worthwhile to focus on the individual needs of patients who require psychosocial support because of their cancer diagnosis and relevant risk factors such as poverty, rather than because of the pandemic.

## Data Availability

The data underlying this article are available online at the Open Science Framework (OSF).

https://doi.org/10.17605/OSF.IO/7THFY

## Notes

### Author Contributions

Conceptualization: ELZ, AG

Data curation: ELZ, CS

Formal analysis: ELZ, MK, JP

Investigation: ELZ, CS, MU, TF, RB, EKM, FA, LK, BK, SZM, MR, MTK, PBS, PV

Methodology: ELZ, MK, AG

Project administration: ELZ, SL, AG

Validation: ELZ, MK, JP

Visualization: ELZ

Supervision: AG

Writing - original draft: ELZ, MK

Writing - review & editing: CS, JP, SL, MU, TF, RB, EKM, FA, LK, BK, SZM, MR, MTK, PBS, PV, AG

### Data Availability Statement

The data underlying this article are available in the Open Science Framework (OSF), at https://doi.org/10.17605/OSF.IO/7THFY

## References

1. Labaki C, Peters S, Choueiri TK. Treatment Decisions for Patients with Cancer during the COVID-19 Pandemic. Cancer Discovery. 2021;11(6):1330–1335. doi:10.1158/2159-8290.CD-21-0210

2. Rezvi MR, Tonmoy MdSB, Khan B. The mental health of adolescents following the COVID-19 pandemic in Bangladesh. Asian Journal of Psychiatry. 2022;78:103309. doi:10.1016/j.ajp.2022.103309

3. Zeilinger EL, Oppenauer C, Knefel M, et al. Prevalence of anxiety and depression in people with different types of cancer or haematologic malignancies: a cross-sectional study. Epidemiology and Psychiatric Sciences. 2022;31:e74. doi:10.1017/S2045796022000592

4. Gaiger A, Lubowitzki S, Krammer K, et al. The cancer survival index—A prognostic score integrating psychosocial and biological factors in patients diagnosed with cancer or haematologic malignancies. Cancer Medicine. 2022;11(18):3387–3396. doi:10.1002/cam4.4697

5. Unseld M, Zeilinger EL, Fellinger M, et al. Prevalence of pain and its association with symptoms of post-traumatic stress disorder, depression, anxiety and distress in 846 cancer patients: A cross sectional study. Psychooncology. 2021;30(4):504–510. doi:10.1002/pon.5595

6. Islam JY, Vidot DC, Camacho-Rivera M. Evaluating mental health–related symptoms among cancer survivors during the COVID-19 pandemic: An analysis of the COVID impact survey. JCO Oncol Pract. 2021;17(9):e1258–e1269. doi:10.1200/OP.20.00752

7. van de Haar J, Hoes LR, Coles CE, et al. Caring for patients with cancer in the COVID-19 era. Nat Med. 2020;26(5):665–671. doi:10.1038/s41591-020-0874-8

8. Ayubi E, Bashirian S, Khazaei S. Depression and Anxiety Among Patients with Cancer During COVID-19 Pandemic: A Systematic Review and Meta-analysis. J Gastrointest Canc. 2021;52(2):499–507. doi:10.1007/s12029-021-00643-9

9. Ernst M, Beutel ME, Brähler E. Cancer as a risk factor for distress and its interactions with sociodemographic variables in the context of the first wave of the COVID-19 pandemic in Germany. Sci Rep. 2022;12(1):2021. doi:10.1038/s41598-022-06016-x

10. van de Poll-Franse LV, de Rooij BH, Horevoorts NJE, et al. Perceived care and well-being of patients with cancer and matched norm participants in the COVID-19 crisis: Results of a survey of participants in the Dutch PROFILES registry. JAMA Oncol. 2021;7(2):279–284. doi:10.1001/jamaoncol.2020.6093

11. Rentscher KE, Zhou X, Small BJ, et al. Loneliness and mental health during the COVID-19 pandemic in older breast cancer survivors and noncancer controls. Cancer. 2021;127(19):3671–3679. doi:10.1002/cncr.33687

12. Serrano-Alarcón M, Kentikelenis A, Mckee M, Stuckler D. Impact of COVID-19 lockdowns on mental health: Evidence from a quasi-natural experiment in England and Scotland. Health Economics. 2022;31(2):284–296. doi:10.1002/hec.4453

13. Zeilinger EL, Lubowitzki S, Unseld M, et al. The impact of COVID-19 on cancer care of outpatients with low socioeconomic status. Int J Cancer. 2022;151(1):77–82. doi:10.1002/ijc.33960

14. Elm E von, Altman DG, Egger M, Pocock SJ, Gøtzsche PC, Vandenbroucke JP. The Strengthening the Reporting of Observational Studies in Epidemiology (STROBE) statement: guidelines for reporting observational studies. The Lancet. 2007;370(9596):1453–1457. doi:10.1016/S0140-6736(07)61602-X

15. Zigmond AS, Snaith RP. The Hospital Anxiety and Depression Scale. Acta Psychiatr Scand. 1983;67(6):361–370. doi:10.1111/j.1600-0447.1983.tb09716.x

16. Stoll C, Kapfhammer HP, Rothenhäusler HB, et al. Sensitivity and specificity of a screening test to document traumatic experiences and to diagnose post-traumatic stress disorder in ARDS patients after intensive care treatment. Intensive Care Med. 1999;25(7):697–704. doi:10.1007/s001340050932

17. Zeilinger EL, Nader IW, Wiedermann W, et al. Latent structure and measurement invariance of the Hospital Anxiety and Depression Scale in cancer outpatients. International Journal of Clinical and Health Psychology. 2022;22(3):100315. doi:10.1016/j.ijchp.2022.100315

18. Temel JS, Pirl WF, Recklitis CJ, Cashavelly B, Lynch TJ. Feasibility and validity of a one-item fatigue screen in a thoracic oncology clinic. Journal of Thoracic Oncology. 2006;1(5):454–459. doi:10.1016/S1556-0864(15)31611-7

19. Grant S, Aitchison T, Henderson E, et al. A comparison of the reproducibility and the sensitivity to change of visual analogue scales, Borg scales, and Likert scales in normal subjects during submaximal exercise. Chest. 1999;116(5):1208–1217. doi:10.1378/chest.116.5.1208

20. Statista. Austria - Poverty Thresholds According to Household Types 2021 [Österreich - Armutsgrenze nach Haushaltstypen 2021]. Statista. Published 2022. Accessed November 9, 2022. https://de.statista.com/statistik/daten/studie/1270720/umfrage/armutsgrenze-in-oesterreich-nach-haushaltstypen/

21. Hammouri HM, Sabo RT, Alsaadawi R, Kheirallah KA. Handling skewed data: A comparison of two popular methods. Applied Sciences. 2020;10(18):6247. doi:10.3390/app10186247

22. Rubin M. When to adjust alpha during multiple testing: a consideration of disjunction, conjunction, and individual testing. Synthese. 2021;199(3):10969–11000. doi:10.1007/s11229-021-03276-4

23. Zeilinger EL, Gaiger A. Data from: Anxiety and depression in cancer outpatients. OSF. Published online 2022. doi:10.17605/OSF.IO/7THFY

24. Amram O, Robison J, Amiri S, Pflugeisen B, Roll J, Monsivais P. Socioeconomic and Racial Inequities in Breast Cancer Screening During the COVID-19 Pandemic in Washington State. JAMA Network Open. 2021;4(5):e2110946. doi:10.1001/jamanetworkopen.2021.10946

